# A Method to Redesign and Simplify Schedules of Assessment and Quantify the Impacts. Applications to Merck Protocols

**DOI:** 10.1101/2024.01.02.24300749

**Authors:** Steven R. Cummings, Scott Chetham, Andy Lee

**Affiliations:** San Francisco Coordinating Center, California Pacific Medical Center Research Institute, Mission Hall: 550-16th Street, 2nd Floor, San Francisco, CA 94143; Department of Epidemiology and Biostatistics, University of California, San Francisco, 550-16th Street, 2nd Floor, San Francisco, CA 94143; Faro Health Inc., 3366 N Torrey Pines CT, Suite 225. La Jolla, CA 92037; Global Clinical Trial Operations, Merck & Co., Inc., Rahway, NJ, USA

## Abstract

The growing complexity of biopharmaceutical sponsored trials has adverse impacts on increased burdens on participants, clinical sites, and sponsors, including greater difficulty recruiting and retaining participants, difficulty engaging sites to participate in trials, excessive cost of trials, and increased cycle times. The schedule of assessments (SoAs) is the origin of and blueprint for complexity that is often generated by copying and pasting from previous SoAs. We developed an approach, termed Lean Design, for redesigning SoAs, restarting SoAs from ‘ground zero’, challenging the addition of assessments using several principles of trial design. We employed a system, the Faro Trial Designer Tool, to quantify the impacts of changes in an SoA to provide real-time feedback to the team and sponsor. We applied the approach in workshops with teams for six clinical trials in various stages of design and implementation. The approach resulted in recommendation for substantial savings in participant and site staff time, costs, and complexity of the trials. Application of this approach to very early stages of protocol design has the potential to reduce the complexity of biopharmaceutical sponsored trials and its consequences.

## Introduction

Industry-sponsored clinical trials have become excessively complex. The growing amount of data to collect, monitor, clean, process, analyze, interpret, and review by regulators has also contributed to increases in the mean duration of both Phase II and Phase III clinical trials [1]. The burden of trials to participants is increasing the difficulty of recruiting and retaining participants. The burden is greatest for people in lower socioeconomic and minority groups with inflexible job schedules and lack of childcare. Burden on staff diminishes the attractiveness of trials to clinical sites.

ICH guideline E19 encourages sponsors, to reduce the amount data collected and submitted for review that have liittle or no relevance to the molecule’s indications and plausible adverse effects.[1] However, little progress has been made in reducing the complexity in protocol design and fundamentally new approaches are required.[2]

Teams that design protocols get remarkably little feedback about the impacts of their choices on the time required of people to participate and the complexity of the trial for staff, or the cost of elements included in the trial. Thy see no data about the impact of collecting assessments to. Those metrics are also valuable to the company to monitor the cost and impacts of protocols and to consider ways to reduce them. Faro Health Inc., (www.farohealth.com) developed a tool that quantifies all key aspects of a trial that reveals the impact of choices made in the design of trial protocols.

A major source of increasing complexity is reliance on the process of “Copy-and-Paste.” The most efficient approach to creating a schedule of assessment is to copy one from previous trials or a standard template. Further development of the SoA typically focuses on selection of primary and secondary endpoints and assessments that might provide advantages over competitors. Some are added to anticipate questions from regulatory agencies.

Reviews of protocols consider the likelihood of successful registration, safety, and implications for marketing. The complexity of the SoAs is uncommonly a focus of reviews. The SoAs that emerge with additions, become templated for other trials. Complexity snowballs without constraints.

A few studies of the costs of clinical trials have shown that predictable factors including the number of participants, sites, the duration of trials, all influence the cost of trials.[3,4] [5,6] No study has measured the contributions of inefficiencies of protocol design to the cost of trials and none have estimated the impact of inefficient trial design on the hours required by participants to participate in a trial and clinical staff to conduct them. Most importantly, no study has demonstrated how much, reduction in unnecessary features of schedules of assessment may reduce burdens and costs. Merck has undertaken a project, termed ‘Lean Design’ to simplify trials and quantify the potential impacts.

## Methods

### Lean Design

Simplifying SoAs stops the copy-and-paste habit. It begins with an SoA (Ground Zero) that includes only the primary aim with its sample size and duration of follow-up but is otherwise blank. Additions are considered according to fundamental principles that encourage simplification, summarized in Table 1.

**Table 1.**
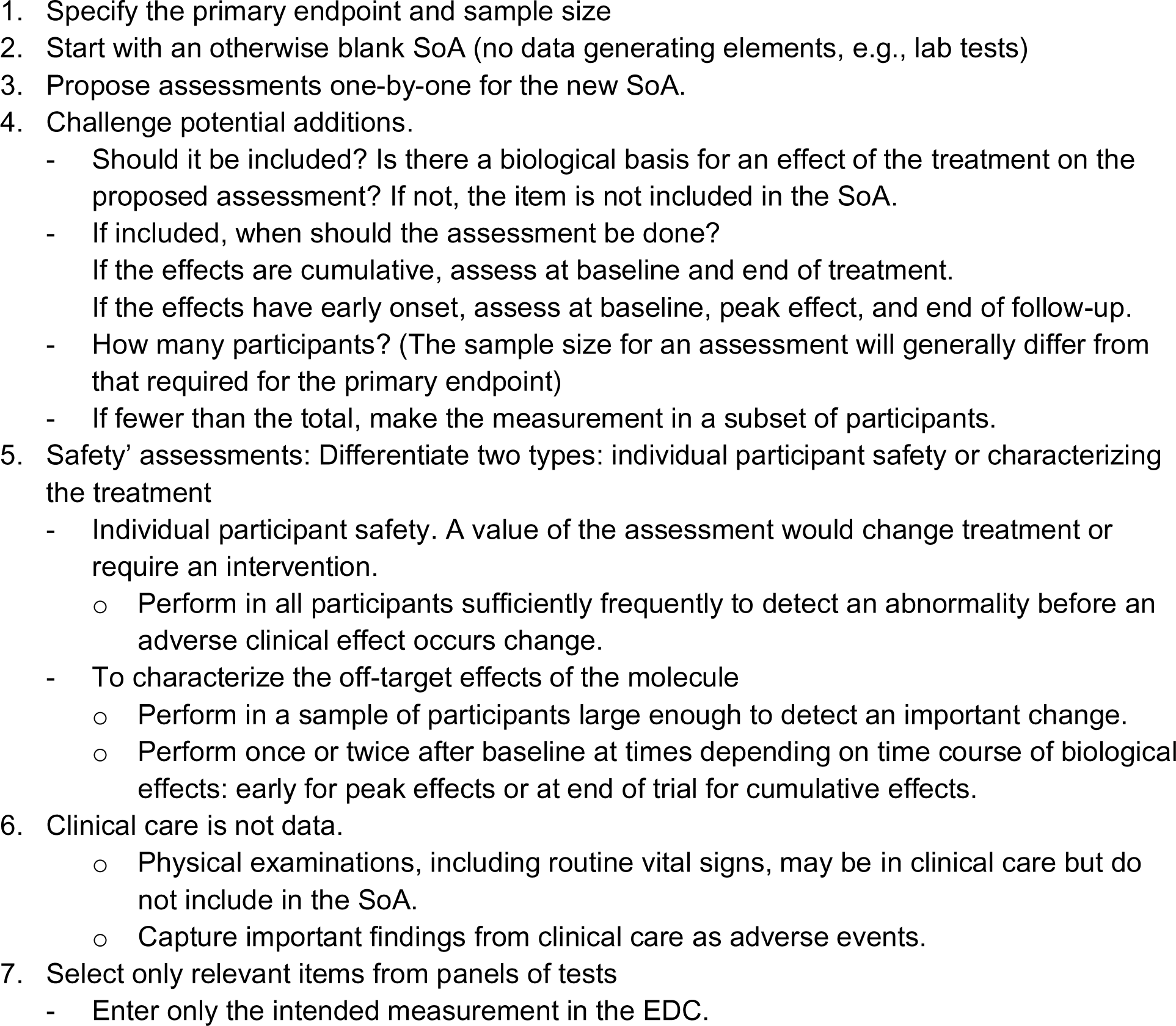
Selected Lean Design Process and Principles for Developing Schedules of Activities.

### Faro Smart Design Tool

The Faro Smart Design Tool (Faro Health Inc.) estimates the hours, burden, types of resources needed, costs, and complexity of the SoA. It can provide *real-time* estimates of the patient burden, patient visit times, required site staff time, activity cost, blood volumes and operational complexity for a site. The Faro Smart Designer software can estimate these metrics in real-time to see how each assessment contributions to the burden of executing the trial.

### Application to specific protocols

Merck clinical trial leadership selected six protocols from different therapeutic areas for Lean Design workshops: cardiovascular disease and other medical conditions, psychiatry and cancer. Initial workshops oriented the team to the Lean Design method. These were followed by an in-person workshops that reviewed the SoA using the Lean Design principles. Workshops were led by an expert in clinical trial design and also a member of the Faro team.

Teams were told that the workshops were exercises and changes recommended in workshops were not mandatory. Most protocols were in late stages of development and approval which precluded making many of the changes that were recommended or agreed to in the workshops.

The workshops focused on the SoA. Administrative and essential procedures, such as the administration of the study drug, were included in the SoA but not in the design exercise. The primary endpoint and collection of adverse events were retained, although the frequency and timing or the assessments were examined. There were no changes proposed to the overall sample size or the plan for collection of AEs. However, timepoints for assessments were removed from the SoA. This generated a SoA that included only these essential administrative elements (Ground Zero).

Study teams proposed specific assessments for the SoA. Each was considered in accordance with Lean Design Principles (Table 1). First, potential additions were challenged as to whether they were plausible biological effects of the investigational on an assessment. A proposal to include a complete blood count was considered based on plausible effects of the treatment on any components of the blood count. Second, if there was a biological basis for an effect, then the timing and frequency of the measurements was considered. If the drug had a rapid onset of effect(s) that remained constant, then a measurement at the time of anticipated peak effect was included and the rationale for subsequent measurements was questioned. If the drug was expected to have cumulative effect(s), for example, effect on the progression of fibrotic changes on imaging, it was recommended that the assessment would be done at baseline and once at the end of treatment or of the observation period.

Some assessments can be done in subsamples of participants. The sample size for a trial is based on the primary endpoint and may not be relevant to other assessments. For large trials, assessments in subsets were proposed for some measurements. Appropriate sample size requires judgement about the minimal size of the effect that would be important to detect. The subsamples could be of first participants or in a few clinical sites.

Two types of safety assessments were identified. First, assessments for individual participant safety that would need to be done periodically in all participants to discover actionable abnormalities. For example, for a drug with treatment of psychiatric effects that might increase the risk of suicide, then the development of a suicidal ideation would be made in all participants frequently enough to detect suicidal ideations before a suicide attempt. Second, all other assessments would be done to characterize the profile of adverse effects. It was noted that these could be done in limited sample sizes sufficient to detect important effects and performed just at times consistent with the time course of the biological effects of the treatment.

Assessments for routine clinical care, such as physical exams, are not included in the SoA. They may be part of the protocol but, rather than including them as data in the SoA, abnormalities they discover can be reported as adverse events. Some physical examinations collect essential data, such as examination of sites of cancer recurrence to assess progression-free survival.

When panels of measurements are proposed, such as a chemistry panel, the element(s) that might be affected by the drug, are entered as data, but other elements were not entered into the study database. This avoids substantial downstream costs for trial monitoring, reconciliation of data by site personnel, follow-up of out-of-range data not relevant to the treatment, and data analyses and reporting to the FDA.

The workshop leaders pushed back on several common reasons for including activities that were not allowed in the construction of new SoAs. These reasons included, the activity is a standard part of protocols; the participant is already at the clinical site, and the assessment is part of routine medical care for the condition, or a sample has already been obtained (for a different purpose). The same principles were applied to laboratory and physical measurements and questionnaires or interviews, for example, for patient-reported outcomes (PRO). Most PROs are collected in all trial participants without consideration of intervention effect, or sample size. The workshop leaders also challenged the inclusion of PK/PD sampling in all participants at multiple visits, without a sample size rationale.

Study teams had. “homework” follow-up from the workshop, study teams met to discuss recommendations from the workshop, and which changes they would adopt. In a follow-up meeting, the teams reported their revised version of the SoA to the leaders of the Lean Design workshop. Reasons for retaining assessments that had not been recommended in the Workshop were discussed.

The Faro team quantified the changes in the impacts of the SoA, comparing the original to a version that represented the recommendations and proposed changes made during the workshop. Using the tool and extensive databases of hours, direct activity costs, and complexity of the three versions and differences between the original, the ‘Workshop,’ and the version with changes that the team adopted (Tables 2 - 5).

**Table 2.** Impact of Lean Design workshops on patient hours.

| Therapeutic Areas<br>Number of<br>Participants.<br>Duration of Trials<br>(wks.) | Baseline and Change in Patient Hours (hrs.) |  |  |  |  |  |
| --- | --- | --- | --- | --- | --- | --- |
|  | Per Participant |  |  | Total for Trial |  |  |
|  | Pre<br>Workshop | Result of<br>Workshop | Adopted | Pre<br>Workshop | Result of<br>Workshop | Adopted |
| Liver Disease<br>N = 328, 78 wks. | 40.4 | -15.10 | -3.7 | 13,251 | -4,953 | -1,213 |
| Cardiac Disease<br>N = 12600, 250 wks. | 47.8 | -9.4 | -4.3 | 602,280 | -118,440 | -54,180 |
| Pulmonary Disease<br>N = 164, 122 wks. | 395.2 | -47.2 | -9.3 | 64,813 | -7,734 | -1,525 |
| Cancer<br>N = 1170, 128 wks. | 168.5 | -6.0 | 0.0 | 197,145 | -7,020 | 0 |
| Infectious Disease<br>N = 2596, 24 wks. | 24.3 | -4.0 | -4.8 | 63,083 | -16,932 | -12,461 |
| Psychiatric Disease<br>N = 500, 15 wks. | 45.4 | -7.9 | -5.8 | 22,700 | -4,040 | -2,900 |

**Table 3.** Impact of Lean Design workshops on hours spent by site staff.

| Therapeutic Areas<br>Number of Participants.<br>Duration of Trials (wks.) | Baseline and Change in Site Staff (hrs.) |  |  |  |  |  |
| --- | --- | --- | --- | --- | --- | --- |
|  | Per Participant |  |  | Total for Trial |  |  |
|  | Pre<br>Workshop | Result of<br>Workshop | Adopted | Pre<br>Workshop | Result of<br>Workshop | Adopted |
| Liver Disease<br>N = 328, 78 wks. | 60.70 | -29.10 | -3.70 | 19,910 | -9,545 | -1,214 |
| Cardiac Disease<br>N = 12600, 250 wks. | 68.30 | -9.10 | -4.80 | 860,580 | -114,660 | -60,480 |
| Pulmonary Disease<br>N = 164, 122 wks. | 108.80 | -46.30 | -2.10 | 17,843 | -7,588 | -344 |
| Cancer<br>N = 1170, 128 wks. | 195.90 | -6.00 | 0.00 | 229,203 | -7,020 | 0 |
| Infectious Disease<br>N = 2596, 24 wks. | 23.20 | -3.40 | -3.90 | 60,227 | -13,537 | -10,124 |
| Psychiatric Disease<br>N = 500, 15 wks. | 76.90 | -10.10 | -7.60 | 38,450 | -5,200 | -3,800 |

**Table 4.** Impact of Lean. Design workshops on changes in direct activity costs of the trial.

| Therapeutic Areas<br>Number of Participants.<br>Duration of Trials (wks.) | Baseline and Change in<br>Direct Activity Cost |  |  |  |  |  |
| --- | --- | --- | --- | --- | --- | --- |
|  | Per Participant |  |  | Total for Trial **(,000)** |  |  |
|  | Pre<br>Workshop | Result of<br>Workshop | Adopted | Pre<br>Workshop | Result of<br>Workshop | Adopted |
| Liver Disease<br>N = 328, 78 wks. | \$38,000 | -\$9,000 | -\$2,000 | \$12,333 | -\$2,821 | -\$394 |
| Cardiac Disease<br>N = 12600, 250 wks. | \$44,000 | -\$9,000 | -\$4,000 | \$556,920 | -\$119,700 | -\$57,960 |
| Pulmonary Disease<br>N = 164, 122 wks. | \$345,000 | -\$28,000 | \$0 | \$56,516 | -\$4,595 | \$0 |
| Cancer<br>N = 1170, 128 wks. | \$173,000 | \$0 | \$0 | \$202,176 | \$0 | \$0 |
| Infectious Disease<br>N = 2596, 24 wks. | \$16,000 | -\$3,000 | -\$2,000 | \$41,796 | -\$7,179 | -\$6,231 |
| Psychiatric Disease<br>N = 500, 15 wks. | \$30,000 | -\$4,000 | -\$1,000 | \$15,150 | -\$1,993 | -\$450 |

**Table 5.** Impact of Lean Design workshops on complexity of the trial.

| Therapeutic Areas<br>Number of Participants.<br>Duration of Trials (wks.) | Baseline and Change in Complexity (RVU) |  |  |  |  |  |
| --- | --- | --- | --- | --- | --- | --- |
|  | Per Participant |  |  | Total for Trial |  |  |
|  | Pre<br>Workshop | Result of<br>Workshop | Adopted | Pre<br>Workshop | Result of<br>Workshop | Adopted |
| Liver Disease<br>N = 328, 78 wks. | 55.22 | -14.93 | 0.00 | 55.22 | -14.93 | 0.00 |
| Cardiac Disease<br>N = 12600, 250 wks. | 72.18 | -12.49 | -7.64 | 72.18 | -12.49 | -7.64 |
| Pulmonary Disease<br>N = 164, 122 wks. | 444.79 | -150.48 | -41.71 | 444.79 | -150.48 | -41.71 |
| Cancer<br>N = 1170, 128 wks. | 858.79 | 0.00 | 0.00 | 858.79 | 0.00 | 0.00 |
| Infectious Disease<br>N = 2596, 24 wks. | 112.65 | -18.38 | -29.86 | 112.65 | -18.38 | -29.86 |
| Psychiatric Disease<br>N = 500, 15 wks. | 77.74 | -0.07 | -1.12 | 77.74 | -0.07 | -1.12 |

## Results

The Lean Design workshops usually generated major potential changes in SoAs that would result in substantial savings in participant and staff time, costs, and complexity (Tables 2-4). Accumulated across numerous trials in a sponsor’s portfolio, durable effects of the Lean Design method would have substantial effects on the burdens and costs and cycle times of a sponsor’s clinical trials.

Common changes from the original version of the SoA included fewer assessments, such as specific laboratory tests. Changes in the number and timing of some assessments were also made. For example, a CBC done to screen for acute off-target effects of the drug were recommended to be included only at baseline and one or two early timepoints. The realization that clinical care is not always useful clinical trial data commonly led to deletion or substantial reduction in the frequency of physical examinations with specification that only abnormalities would be collected as adverse events. In a few SoAs for large trials, a few assessments done to characterize the effects of the treatment were planned for smaller subsets of participants, generally done early in follow-up to inform whether there were any effects that needed continued collection.

The number and amounts of changes varied considerably across the trials (Tables 2-4). Recommendations would have reduced participant hours by 4 to 47 per participant per trial. The total changes in hours and costs for the trial were, as expected, proportional to the size and duration of the trial. For example, the $9,000 recommended reduction per participant in the cardiovascular trial with 12,600 participants followed for 250 weeks would save nearly $120 million for the whole trial. Few changes were recommended for the oncology trial. This reflected the highly standardized approaches to assess cancer progression in oncology trials.

Most study teams adopted some but not all of the recommended revisions. This was expected because the workshops were considered exercises whose results were not mandatory. Adoption of changes was often limited because of the late stage of the SoAs that were reviewed. Even when there was no plausible biological reason that a treatment would influence a laboratory test or previous data showing no effect, some study teams were not comfortable with the uncertainty that some unexpected abnormal result might arise. Additionally, when asked, the quality of life and pharmacology groups simply asserted that no changes could be made in the number, frequency or sample size of patient reported outcomes or PK-PD assessments.

Teams sometimes raised concerns about the potential importance of data for the FDA or other regulatory bodies. In general, they tried to anticipate FDA interests by including more assessments. When the agency did not comment on the assessments, the team assumed that the assessments had been approved and could not be changed. However, it was pointed out that ICH guidelines have recommended reductions in the amount of data collected in trials [1]. Additionally, teams may be better served by proposing a very lean version of the protocol and SoA and then adding elements back in if required by the agency. The items that are required by the FDA may reveal issues that have arisen in competitors’ trials.

## Discussion

There is a growing recognition that the increasing protocol complexity is unsustainable. The Lean Design workshops identified and recommended major changes in most of the SoAs that would substantially reduce participant and staff hours, costs and complexity of the trials.

Workshops were described as exercises and that recommended changes were not mandatory. Protocols were generally in the late stages of development and approval, so many recommendations were not adopted. However, changes that were adopted would have resulted in substantial changes in the impacts of the revised SoA. For example, in the cardiovascular trial, adopting only $4,000 of the $9,000 recommended reductions in cost per participant would reduce the total cost of the trial by almost $58 million. Even when changes from the workshop were not adopted, team members often said that they agreed with the approach and the principles would inform their future development of SoAs.

More of the substantial changes identified in workshops would have been adopted if protocols had undergone the Lean Design exercise earlier in their development, ideally when SoAs were initially drafted. Nevertheless, for most protocols, even the more limited adoption resulted in savings of many hours and costs. Feedback about the impact of design choices, provided by the Faro Smart Designer software, revealed the impacts of choices in designing the protocol to influence choices about the inclusion or timing of assessments. It is likely to have the greatest impact when used in real time from the start of protocol design.

Importantly, the estimated costs and savings from the SoA pertained only to items listed in the SoA. They did not account for the fully loaded cost to the sponsor, including data collection, monitoring, cleaning, reconciliation, analysis, and reporting. Similarly, the ‘downstream’ impacts on third party and other sponsor activities were not assessed. This is an important consideration for panels of laboratory tests done for the purpose of assessing one or few of dozens of elements. The costs of monitoring and query resolution is also an important burden for clinic staff and sponsors. A comprehensive database of downstream costs of excess data points capture is in development.

There are various approaches to controlling the complexity of trials. Review committees generally focus on major issues such as regulatory approval and competitive position. Some sponsors may try SoA templates, algorithms, or artificial intelligence to identify elements that are not essential. Artificial intelligence is being applied largely to recruitment of and selection, monitoring, and retention of participants.[7–9] There has been no description of its role in designing or improving the efficiency of SoAs. It is not yet clear whether AI could replace medical judgements about the value and patterns of assessments.

The face-to-face approach to rebuilding an SoA in this project may be more effective in making and retaining changes than just presentations about the principles. However, it is impractical for one committee or individual to review all trial protocols under development at a large pharmaceutical company. An approach of training several individuals in a company to apply the methods, armed with a tool that provides feedback about the impacts of choices would facilitate broader effective adoption of Lean Design and its associated principles.

Simplification of trails across a company requires support and promotion from the leadership of the company and therapeutic areas. It requires buy-in from groups that can enable simplification including regulatory, clinical trial operations, and biostatistics groups. It is important to have analyses of the impacts of changes from a simplification program to support implementation of the process.

Propelled by the results of the Lean Design workshops, Merck launched a company-wide project to implement its principles across all therapeutic areas. Simplification, including lean design principles, is considered in the initial design SoAs. Leaders from therapeutic areas are trained in the Lean Design principles to extend the effects to many trials in all therapeutic areas.

While the value of the Lean Design process has been demonstrated in one very large pharmaceutical company; the application and results may differ in other companies. The generalizability of the approach and results should be tested in other companies.

## Conclusion

We conclude that a process of rebuilding protocols from Ground Zero and according to a few principles and supported by quantitative feedback about the impacts of additions, may result in substantial reduction in the number inessential elements included in schedules of assessment with major reductions in the burdens and costs of clinical trials.

## Data Availability

The analyses of the hours and cost savings use a proprietary database and software system. No data had been collected by the protocols analyzed in the report.

## Acknowledgements

The authors would like to acknowledge the work of Laura Galuchie from Merck & Co., Inc., Rahway, NJ, USA for coordinating the Lean Design workshops. We would also like to acknowledge Merck team members from the protocols included in this analysis for their work, along with the Faro Health staff who ran the models.

## Conflicts of Interest

Dr. Cummings is a consultant to Merck & Co., Inc., Rahway, NJ USA and Faro Health, Inc.

Dr. Chetham is Chief Executive Officer of Faro Health Inc.

Mr. Lee is the Head of Global Clinical Trial Operations at Merck & Co., Inc., Rahway, NJ USA.

## Author contributions

Dr. Cummings drafted the manuscript and made critical revisions and prepared it for submission for publication. Dr. Chetham drafted sections of the manuscript, provided the tables of data, critically revised the manuscript, and approved it for submission. Mr. Lee contributed to the initial draft of the manuscript and made critical revisions and approved the final version for submission.

